# Attention-based deep learning for analysis of pathology images and gene expression data in lung squamous premalignant lesions

**DOI:** 10.1101/2025.06.06.25328492

**Authors:** Lingyi Xu, Yohana Kefella, Yichi Zhang, Regan D. Conrad, Kelley E. Anderson, Kostyantyn Krysan, Gang Liu, Erin Kane, Adam Pennycuick, Sam M. Janes, Mary E. Reid, Eric J. Burks, Ehab Billatos, Sarah A. Mazzilli, Vijaya B. Kolachalama, Jennifer E. Beane

## Abstract

Molecular and cellular alterations to the normal pseudostratified columnar bronchial epithelium results in the development of bronchial premalignant lesions representing a spectrum of histology from normal to hyperplasia, metaplasia, dysplasia (mild, moderate, and severe), carcinoma in situ and invasive carcinoma. Several studies have identified molecular alterations associated with lesion histology and progression. The broad and continuous spectrum of histologic and molecular changes makes reproducible stratification of lesions across multiple studies challenging. Here we propose a transformer-based framework that flexibly utilizes transcriptomic and histologic patterns to distinguish lesions with bronchial dysplasia or worse from normal, hyperplasia, and metaplasia. We leveraged H&E whole slide images (WSIs) of endobronchial biopsies and bulk gene expression data (GE) from previously published studies and on-going lung precancer atlas efforts obtained from patients as high-risk for lung cancer. Models trained using both WSIs and GE compared to a single data modality had higher performance. On an external testing dataset of WSIs, the area under the ROC curve (AUROC) of the model trained on WSIs plus GE was 0.761±0.015 compared to 0.690±0.027 for model trained on WSIs. On external testing datasets of GE, the AUROC of the model trained on WSIs plus GE was 0.890±0.023 versus 0.816±0.032 for a model trained on GE. Based on these results, we leveraged data across 4 studies to train a flexible fusion model that allows one or both data modalities to be used in training. The model achieved an AUROC of 0.809±0.036 on external testing WSIs data and 0.903±0.022 on external testing GE data. Despite model training on a binary label, model probabilities are associated with histologic grade and the model identifies gene expression alterations associated with bronchial dysplasia across multiple studies. This framework maps bronchial premalignant lesions that contain at least one data modality into a spectrum of disease. In the future, a framework trained on multiple data modalities may be useful in predicting premalignant disease severity, progression, and interception agent efficacy.

## Introduction

Bronchial premalignant lesions (PMLs) are histologic epithelial abnormalities that are markers of lung cancer risk and precursors to invasive lung squamous cell carcinoma (SCC)^1,2^. Molecular alterations in the normal pseudostratified columnar airway epithelium, driven by poorly understood risk factors including exposure to tobacco smoke, hazardous chemicals, or particulate pollution^3–5^, result in cellular changes that include hyperplasia, metaplasia, dysplasia (mild, moderate, and severe), and carcinoma in situ (CIS) prior to a subset progressing to invasive lung squamous cell carcinoma (SCC). PMLs can be visualized and biopsied via bronchoscopy techniques and pathologic evaluation is conducted using sections of formalin fixed paraffin embedded (FFPE) biopsies stained with hematoxylin and eosin (H&E). Previously, researchers have focused on characterizing transcriptomic, epigenetic, and genetic alterations in PMLs associated with histologic severity and progression to develop methods to identify and treat PMLs with the highest risk of progression to invasive carcinoma^5–11^.

Prior work used biopsy pathology as a reference point to interpret genomic data and define histologic progression in premalignant lung lesions. However, the histologic grading of PMLs is known to have considerable intra- and inter-observer variability, often influenced by whether the reviewing pathologist has pulmonary subspecialty expertise^12^. This variability can limit reproducibility across studies and institutions. To address this variability, our group previously developed a graph perceiver network using digitized whole slide H&E images (WSIs) of endobronchial biopsies to stratify PMLs in the context of normal and lung tumor tissue^13^. While that model showed promise in identifying carcinoma in situ (CIS) lesions with distinct clinical trajectories, it was less effective in resolving lower-grade histologic categories such as hyperplasia and metaplasia. In this study, we focused on dysplastic lesions and presented a transformer-based framework that integrates information from WSIs, and gene expression data obtained from endobronchial biopsies across multiple studies (**Figure 1**). Given the complementary nature of histologic and transcriptomic data, we designed a flexible multimodal learning strategy that can make predictions with one or both modalities present at inference. Such flexibility is critical in real-world clinical and research settings where data completeness varies across cohorts. By enabling cross-study integration and accommodating missing modalities, our approach addresses key limitations in the stratification of early lung lesions and supports more generalizable risk prediction for PMLs.

**Figure 1.**
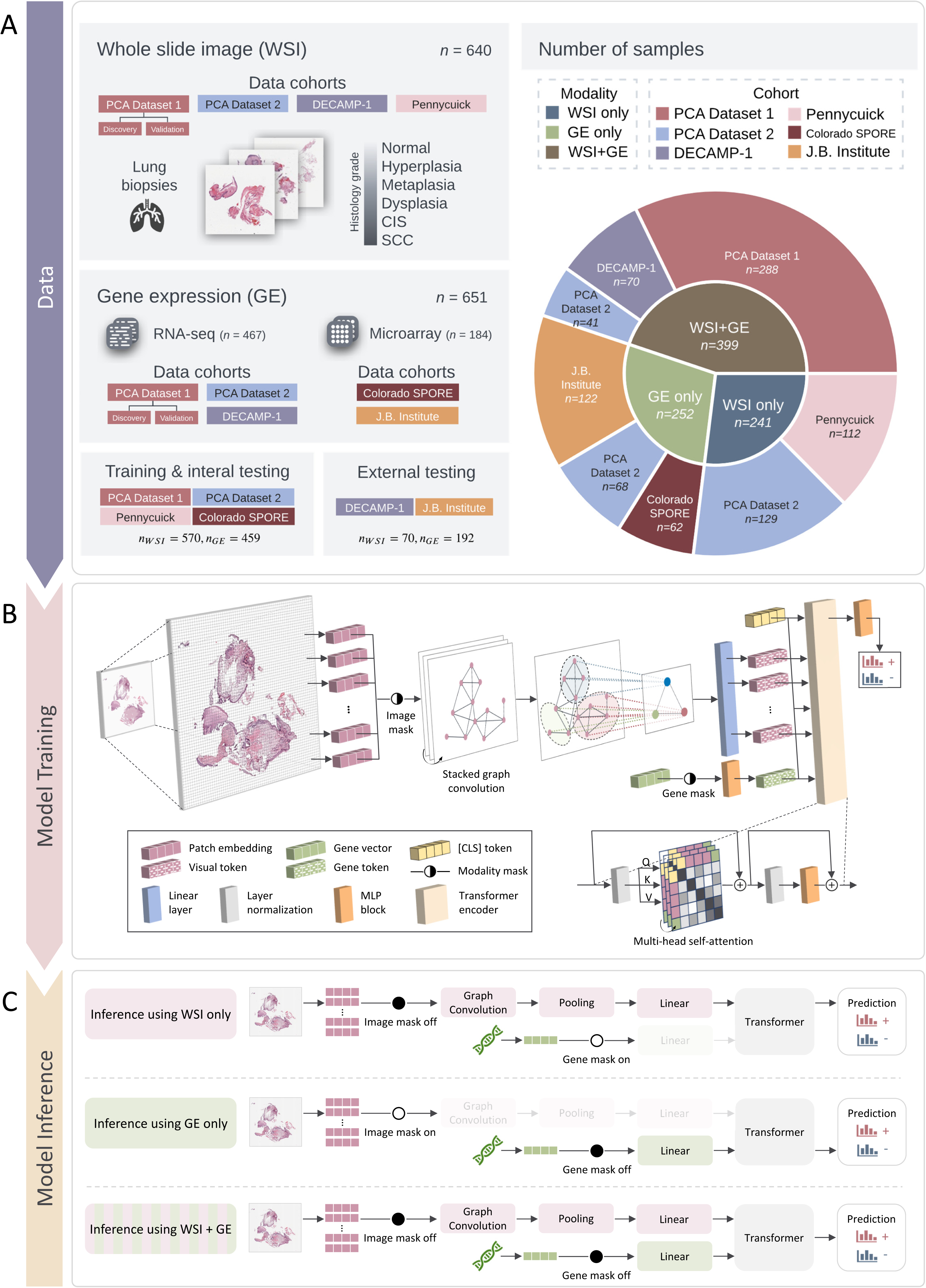
Overview of model architecture, training, and inference. **A.** Our model to classify dysplasia or worse histology versus normal, hyperplasia, or metaplasia (non-dysplasia) was developed on digitized hematoxylin and eosin-stained whole slide images (WSIs) and bulk gene expression data (GE) obtained from endobronchial biopsies aggregated from seven cohorts: PCA Dataset 1 (Discovery and Validation cohorts), PCA Dataset 2, DECAMP-1, Pennycuick, Colorado SPORE, and Jules Bordet Institute. We used samples in the PCA Dataset 1, PCA Dataset 2, Pennycuick, and Colorado SPORE for model training and internal testing. For external testing, we utilized samples from the DECAMP-1 and Jules Bordet Institute cohorts. **B.** A two-branch transformer takes WSIs and GE data as inputs. WSIs were preprocessed into fixed-length vector representations of individual patches. Gene features were preselected based on the association between gene expression and histology. Image and gene masks allow the model to take either single or dual data modalities during training. **C.** The model can perform inference on samples with either WSI or GE or both. With the gene mask on, the model takes the WSI input only, with the image mask on, the model takes the gene input only, and when both masks are off, the model takes both inputs.

**Figure 2.**
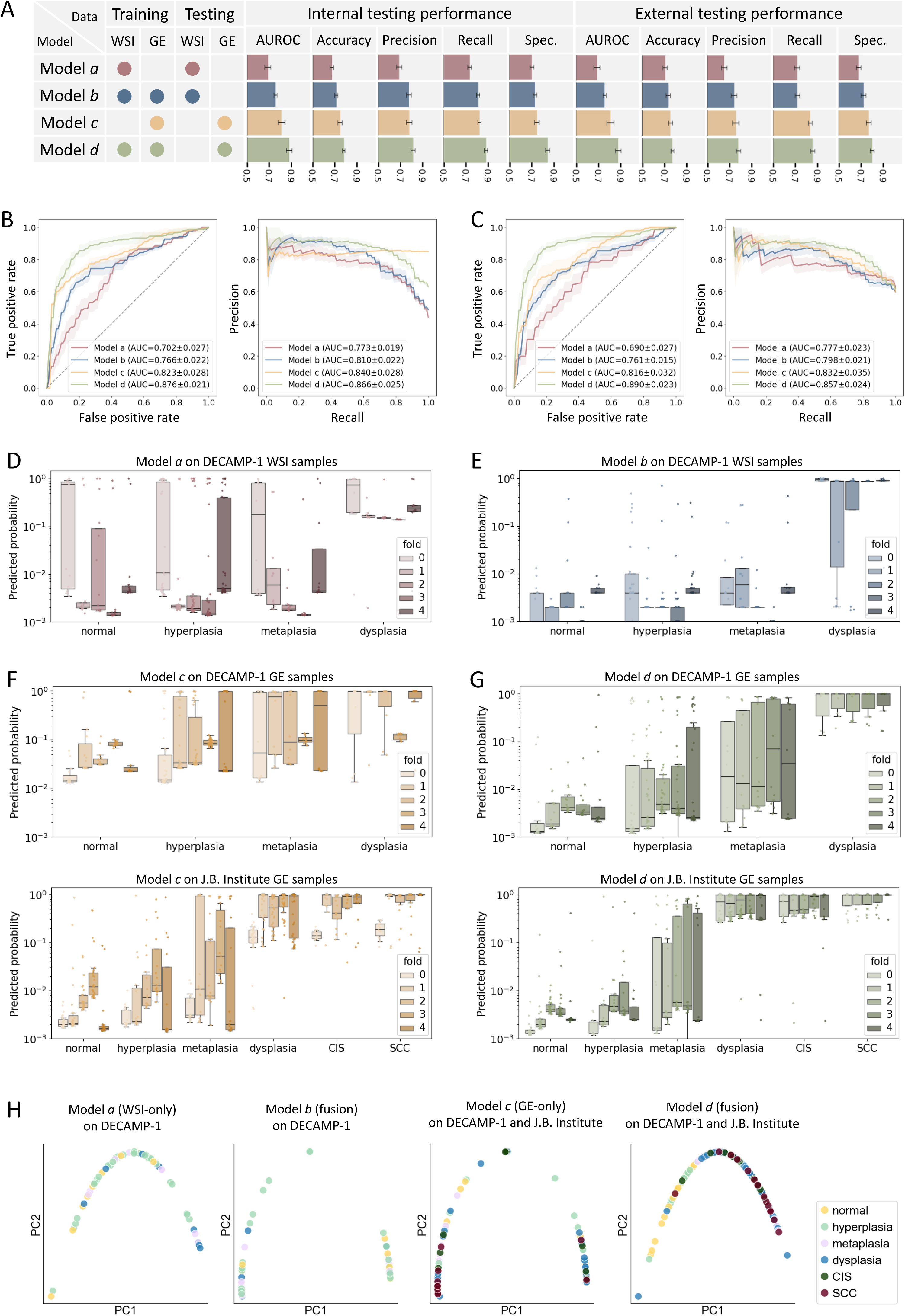
Classification performance of models trained utilizing single versus dual data modalities. **A.** Summary of models and bar charts of model performance across internal and external testing data. Model *a*: trained on WSIs, tested on WSIs; Model *b*: trained on samples with both WSIs and GE, tested on WSIs; Model *c*: trained on GE, tested on GE; Model *d*: trained on samples with both WSI and GE, tested on GE. Performance metrics, including AUROC score, accuracy, precision, recall, and specificity, were visualized in the horizontal bar chart for internal and external testing where the error bars represent the standard deviation across five folds. **B, C.** Receiver operating characteristic (ROC) and precision- recall (PR) curves showing model performance towards binary classification on internal (**B**) and external (**C**) testing samples. The area under the curve (AUC) score was reported with standard deviation across five folds. **D, E, F, G.** External testing probability predictions were visualized with box and strip plots by fold and histology. Models *a* (**D**) and *b* (**E**) were tested on DECAMP-1 WSIs. Model *c* was tested on DECAMP-1 (**F**, top) and J.B. Institute (**F**, bottom) GE samples. Model *d* was also tested on DECAMP-1 (**G**, top) and J.B. Institute (**G**, bottom) GE samples. **H.** PC plots on external testing samples were generated by using the penultimate layer embedding from each model (Model *a* to *d* from left to right). Each point in a PC plot represents a sample with coordinates (PC1, PC2) and the color referring to a specific histologic grade. P-values from the Mann-Whitney U test indicated the statistical significance of PC value distributions between samples with histological grades of dysplasia or worse versus normal, hyperplasia, and metaplasia.

**Figure 3.**
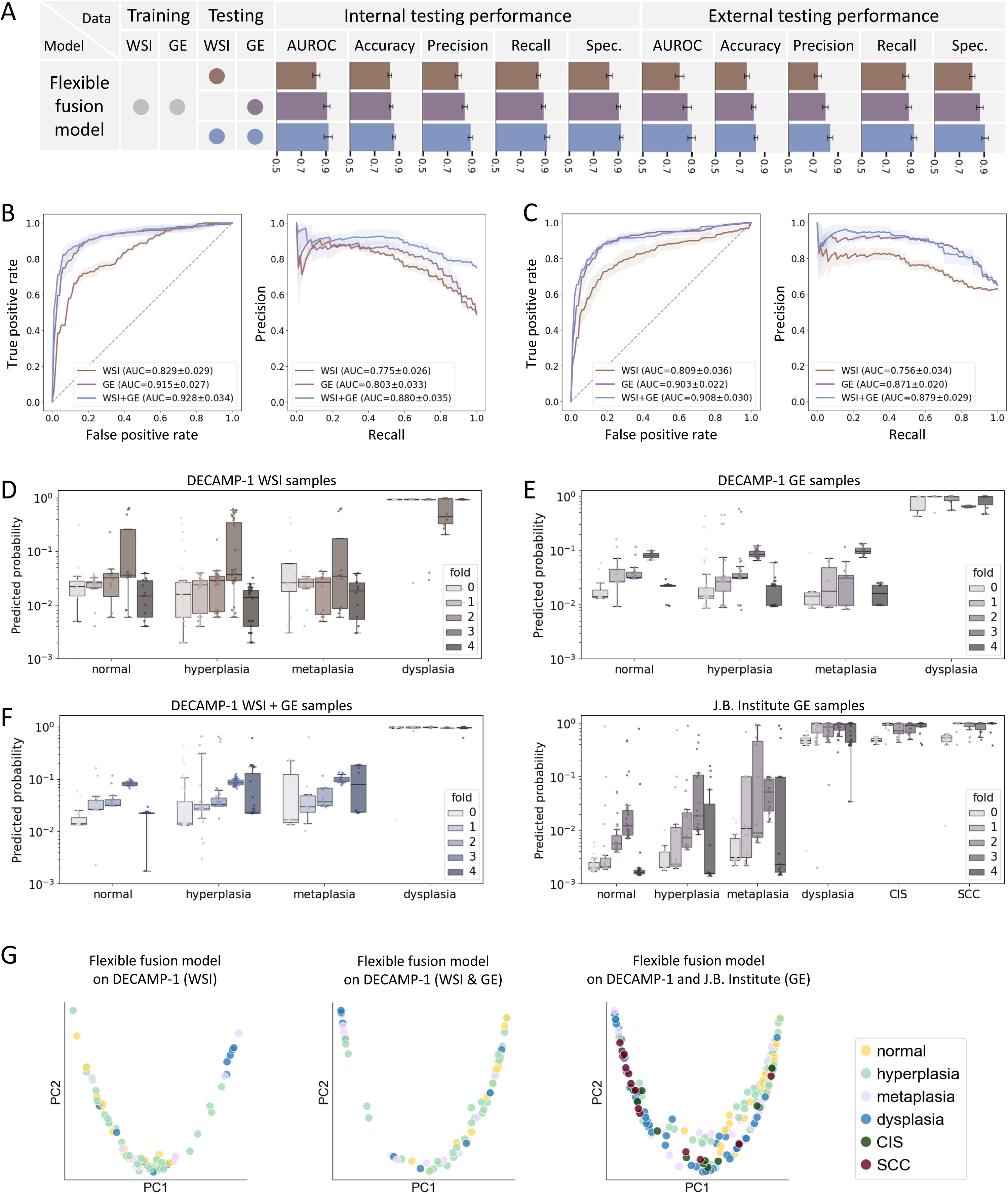
Classification performance of the flexible fusion model. **A.** The flexible fusion model was trained on all samples from training datasets regardless of the number of available data modalities. The flexible fusion model was internally and externally tested on samples with WSI, samples with GE, and samples with both WSI and GE. The same performance metrics as Figure 2 were visually reported in the bar charts. **B, C.** Receiver operating characteristic (ROC) and precision-recall (PR) curves showcase the flexible fusion model performance towards binary classification on internal (**B**) and external (**C**) testing samples (with WSI only, with GE only, or with both WSI and GE). The area under the curve (AUC) score was reported with standard deviation across five folds. **D, E, F.** External testing predictions in probability were visualized with box and strip plots by fold and histology grading, divided into: DECAMP-1 WSI samples (**D**), DECAMP-1 GE samples (**E**, top), J.B. Institute GE samples (**E**, bottom), and DECAMP-1 WSI+GE samples (**F**). **G.** PC plots on external testing samples were generated by using the penultimate layer embedding from the flexible fusion model (from left to right: WSI only samples from DECAMP-1, WSI+GE samples from DECAMP-1, GE samples from DECAMP-1 and J.B. Institute). Each point in a PC plot represents a sample with coordinates (PC1, PC2) and the color referring to a specific histologic grade. P-values from the Mann-Whitney U test indicated the statistical significance of PC value distributions between samples with histological grades of dysplasia or worse versus normal, hyperplasia, and metaplasia.

## Methods

### Study cohorts and data

We obtained H&E stained WSIs and/or bulk gene expression data (GE) from endobronchial biopsies obtained from patients at high risk for developing lung cancer. The histology of the biopsies spans the entire spectrum from normal, hyperplasia, metaplasia, dysplasia (mild, moderate, or severe), CIS, and invasive SCC. All biopsies were assigned the worst histology observed during clinical pathologic evaluation. The goal of our study was to distinguish between normal or low-grade histologic changes (hyperplasia, metaplasia), termed non-dysplasia, and bronchial dysplasia or more advanced lesions (CIS, SCC). The following data was included (**Figure 1A**): 1) Lung Pre-Cancer Atlas (PCA) (Dataset 1: n=179 non-dysplasia, n=109 dysplasia; n=288 WSIs^13^, n=288 GE, GSE109743^6,10^); Dataset 2: n=134 non-dysplasia, n=55 dysplasia, n=37 CIS, n=12 SCC; n=170 WSIs, n=108 GE); 2) Colorado SPORE cohort from Merrick et. al. study^2^ (n=25 non-dysplasia, n=35 dysplasia, n=2 CIS; n=62 GE, GSE114489); 3) Jules Bordet Institute cohort from Mascaux et. al. study^7^ (n=57 non-dysplasia, n=38 dysplasia, n=13 CIS, n=14 SCC; n=122 GE, GSE33479); 4) DECAMP-1 (NCT01785342)^14^ (n=60 non-dysplasia, n=10 dysplasia; n=70 WSIs, n=70 GE); 5) UCLH Surveillance Study from Pennycuick et. al. study^8^ (n=112 CIS; n=112 WSIs, idr0082). We also leveraged Lung PCA bronchial brushings from Dataset 1 and 2 patients that were sampled from normal appearing airway. The brushes were assigned a histologic grade that represented the worst histology among the endobronchial biopsies sampled during the procedure (n=75 non-dysplasia, n=99 dysplasia, 98.6% of patients overlap with Datasets 1 and 2 above). GSE114489 used Affymetrix Human Gene 1.0 ST microarrays, GSE33479 used Agilent Whole Genome microarrays 4*44K G4112F, and all other GE data was obtained via bulk RNA sequencing. PCA Dataset 2 and DECAMP-1 data will be made public upon publication. The pathologic evaluation of PCA Dataset 2 samples was conducted at Roswell Park Comprehensive Cancer Institute or University College London where the samples were collected, and the DECAMP-1 samples were reviewed by Dr. Eric Burks at Boston Medical Center. WSI samples from PCA Dataset 1, PCA Dataset 2, and DECAMP-1 were imaged at 20X magnification. WSI samples from Pennycuick et. al. study were imaged at 40X magnification. Summary statistics and clinical features are reported in **Table 1**.

**Table 1.** Study population of biopsy samples. Whole slide images (WSIs), gene expression data (GE), and corresponding clinical information from six datasets profiling endobronchial biopsies.

| Description | Dataset |  |  |  |  |  |
| --- | --- | --- | --- | --- | --- | --- |
|  | PCA<br>Dataset 1 | PCA<br>Dataset 2 | DECAMP<br>-1 | Pennycuik | Colorado<br>SPORE | J.B.<br>Institute |
| Number of patients | 29 | 52 | 72 | 112 | 62 | 77 |
| Number of WSIs | 288 | 170 | 72 | 112 | NA | NA |
| Number of WSIs per class<sup>1</sup> | 179, 109 | 117, 53 | 62, 10 | 0, 112 | NA | NA |
| Number of patches<sup>2</sup> | 2690, [131, 24226] | 2781, [74, 14190] | 981, [30, 3413] | 763, [62, 5897] | NA | NA |
| Number of RNA-seqs/microarrays | 288 | 108 | 70 | NA | 62 | 122 |
| Number of RNA-seqs/microarrays per class<sup>1</sup> | 179, 109 | 48, 60 | 60, 10 | NA | 25, 37 | 57, 65 |
| Smoking status<sup>3</sup> | 173, 115, 0, 0 | 69, 39, 0, 0 | 38, 33, 1, 0 | 82, 21, 3, 6 | 32, 29, 2, 0 | 67, 55, 0, 0 |
| Age<sup>4</sup> | 36, 190, 50, 12 | NA | 18, 37, 22, 5 | NA | 7, 16, 30, 9 | 11, 44, 34, 33 |
| Gender<sup>5</sup> | 133, 153 | NA | 61, 11 | 94, 18 | NA | 97, 25 |
| Race<sup>6</sup> | 263, 13, 11, 1 | NA | 47, 11, 3, 1 | NA | NA | NA |

<sup>1</sup>Non-dysplasia, Dysplasia or worse histology
<sup>2</sup>Median, Range
<sup>3</sup>Current, Former, Never, Unknown
<sup>4</sup>Binned: 40-49, 50-59, 60-69, 70-79
<sup>5</sup>Males, Females
<sup>6</sup>White, Black or African-American, Asian, Other/unknown

**Table 2.**
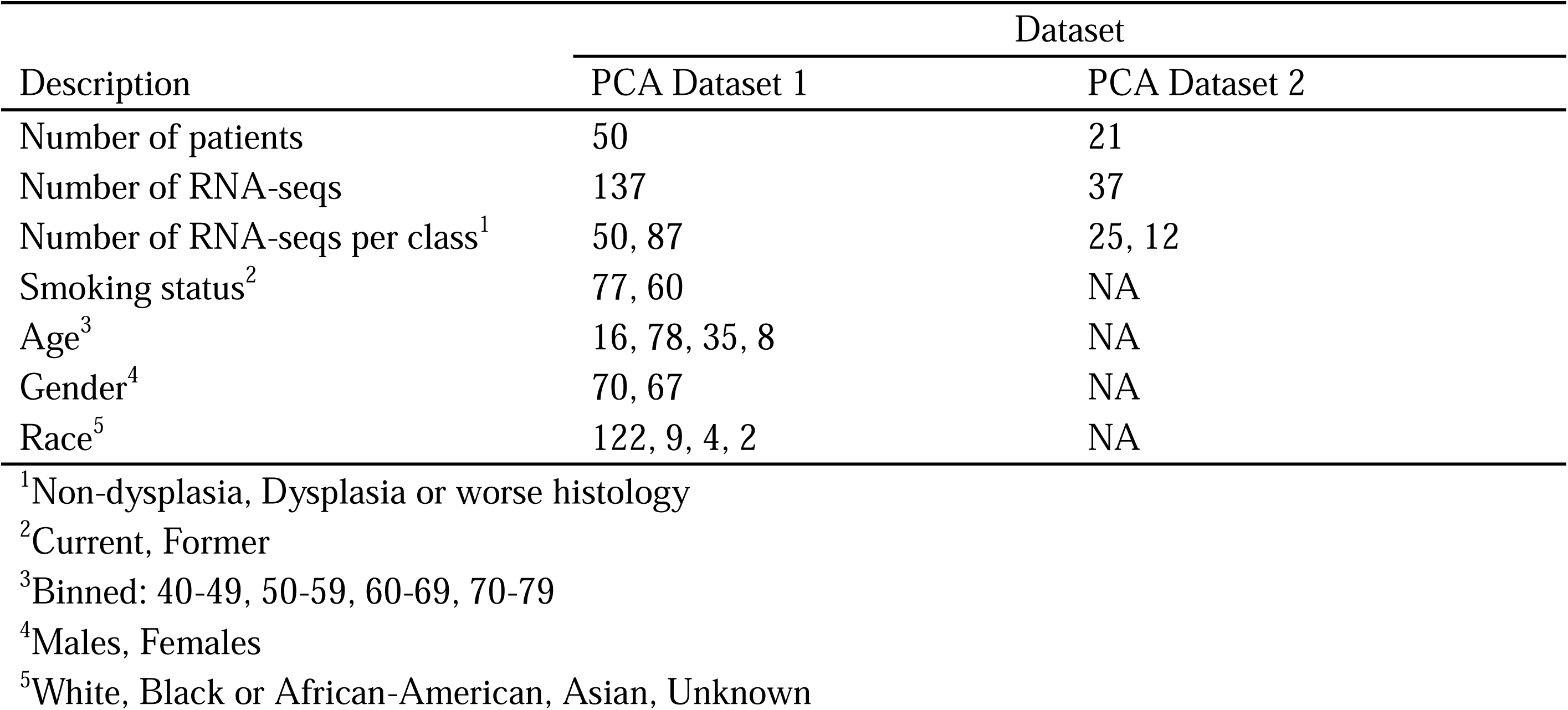
Study population of brushing samples. Gene expression data (GE) and corresponding clinical information from PCA airway brushing samples.

| Description | Dataset |  |
| --- | --- | --- |
|  | PCA Dataset 1 | PCA Dataset 2 |
| Number of patients | 50 | 21 |
| Number of RNA-seqs | 137 | 37 |
| Number of RNA-seqs per class<sup>1</sup> | 50, 87 | 25, 12 |
| Smoking status<sup>2</sup> | 77, 60 | NA |
| Age<sup>3</sup> | 16, 78, 35, 8 | NA |
| Gender<sup>4</sup> | 70, 67 | NA |
| Race<sup>5</sup> | 122, 9, 4, 2 | NA |

<sup>1</sup>Non-dysplasia, Dysplasia or worse histology
<sup>2</sup>Current, Former
<sup>3</sup>Binned: 40-49, 50-59, 60-69, 70-79
<sup>4</sup>Males, Females
<sup>5</sup>White, Black or African-American, Asian, Unknown

### PCA Dataset 2 processing of bulk RNA-seq and H&E WSIs

Endobronchial biopsies from abnormal appearing airway were collected via bronchoscopy from high-risk subjects enrolled in lung cancer screening programs at Roswell Park Comprehensive Cancer Center^6^ and University College London^8^ as previously described. Biopsies obtained for research were either FFPE or flash frozen and embedded in O.C.T. compound. FFPE sections were used for H&E staining and pathologic review and were digitized using the Leica Aperio AT2 scanner at Roswell or the Hamamatsu scanner at UCL. OCT samples were sectioned for research histology review and tissue scrolls were cut for isolation of genomic material. Total RNA was extracted using a minimum of 200 ℒm sections cut on a microtome using the AllPrep DNA/RNA/miRNA Universal Kit (Qiagen). Sequencing libraries were prepared from total RNA samples using Illumina TruSeq Stranded Total RNA Library Prep Gold kit for library preparation. Each sample was sequenced on the Illumina NextSeq 500 or NextSeq 2000 to generate paired-end 100-nucleotide reads. Demultiplexing and creation of FASTQ files were automatically performed using the Illumina BaseSpace platform with default parameters.

### DECAMP-1 processing of bulk RNA-seq and H&E WSIs

Endobronchial biopsies were collected from subjects enrolled in the DECAMP-1 study, “Diagnosis and Surveillance of Indeterminate Pulmonary Nodules (NCT01785342)^14^. Subjects enrolled in DECAMP-1 undergo a baseline bronchoscopy during which two endobronchial biopsies are obtained from 3 predetermined lung sites in the right upper and middle lobes and in the left upper lobe. One biopsy from each site is FFPE, sectioned, and stained with H&E for pathologic review and digitized using the Leica Aperio AT2 scanner at MD Anderson Cancer Center. Total RNA was extracted from selected adjacent frozen biopsies using the AllPrep DNA/RNA/miRNA Universal Kit (Qiagen). Sequencing libraries were prepared from total RNA samples using the Illumina TruSeq Stranded Total RNA Library Prep Gold kit for library preparation. Samples were sequenced on the Illumina HiSeq 2500, the NextSeq500, or the NextSeq 2000 with a target sequencing depth of 40 million 75- or 100-nucleotide paired end reads per sample. Demultiplexing and creation of FASTQ files were automatically performed using the Illumina BaseSpace platform with default parameters.

### Whole slide image pre-processing

WSIs of endobronchial biopsies commonly have multiple sections per slide and artifacts including oil stains, ink marks, and tissue folds (**Figure S1**). Standard WSI segmentation tools, such as CLAM^15^, had difficulty distinguishing tissue from non-tissue areas, often missing important epithelial regions and struggling with slide artifacts. To overcome this, binary masks were used during tessellation to better identify and retain relevant tissue. For every WSI, a binary mask was created by aggregating three hsv color filters: an ink filter to remove blue and green marks, a gray filter to remove black marks and tissue folds, and a saturation and hue filter to remove oil stains. Filter parameters were manually set to achieve a balance between removing artifacts and retaining tissue across WSIs. Artifacts were removed when ink marks, but not oil stains, overlapped the tissues. WSIs were tiled into non-overlapping patches of 128 × 128 pixels, and patches containing less than 15% tissue as indicated by binary mask were excluded.

### Whole slide image graph construction

For each WSI, the high-quality tiled patches were used to construct a graph (**Figure S1**). Every patch was represented as a node (*N*) and a graph was constructed using 8-node adjacency, following the rule that patches are connected if their edges or corners touch. The resulting graph was represented by a feature matrix and an adjacency matrix. The feature matrix of shape *N* × *D* (*D*=768) contained rows of 1 × *D* feature vectors extracted from individual patches through CTransPath^16^, an unsupervised contrastive learning-based feature extraction algorithm pre-trained on The Cancer Genome Atlas (TCGA). The adjacency matrix of shape N × N indicated pairwise connectivity for all nodes in the graph, with elements n =1 when node *i* and *j* are connected.

### Pre-processing of gene expression data

Gene expression matrices for GSE109743^6,10^ and GSE33479^7^ were obtained from NCBI’s GEO^17^. For GSE33479, the Agilent microarray probes contained a mapping to Ensembl transcript IDs. The Ensembl transcript IDs were subsequently mapped to Ensembl Gene IDs using the R package biomaRt^18,19^ and Ensembl^20^ annotation version 113. Multiple probes mapping to a single Ensembl Gene ID were averaged (**Figure S2**). For GSE114489, the Affymetrix CEL files were downloaded from GEO and processed to generate RMA-normalized^21^ data using the Brainarray Ensembl v18.0.0 CDF file^22^. The DECAMP-1 data was aligned using STAR^23^ and hg19. Expression levels of genes were quantitated using RSEM^24^ and Ensembl version 75 annotation. To identify poor quality samples, a principal component analysis of z-score normalized quality control metrics (n=94) computed using RSeQC^25^ and STAR was conducted and samples more than 2 standard deviations from the mean of component 1 were eliminated. The PCA Dataset 2 was processed using the same methods as DECAMP-1 except we utilized our bulk RNA-seq pipeline^26^ on the Terra cloud computing platform that leverages genome version hg38 and Gencode version 34 gene annotation. To identify poor quality samples in the PCA Dataset 2, principal component analyses using z-score normalized quality control metrics or gene expression values were conducted and samples more than 2 standard deviations from the mean of components 1 or 2 were eliminated. Further sample quality checks included 1) identifying samples with low sample-sample correlations based on gene expression (±2 standard deviations from the mean); 2) abnormal heterozygosity rates calculated from Somalier^27^ (±2 standard deviations from the mean); and 3) inconsistencies in Somalier or arcasHLA^28^ outputs between samples derived from the same patient, which may indicate sample swaps or contamination. Samples failing any of these evaluations are also flagged as poor quality and excluded from subsequent analyses. The sample quality filtering was conducted separately for the endobronchial biopsies and bronchial brushings. Gene filtering for all RNA sequencing datasets was conducted as previously described^6^. Log counts per million (cpm) was computed for all RNA sequencing datasets (GSE109743 discovery and validation sets, DECAMP-1, and PCA Dataset 2 endobronchial biopsies, and PCA Dataset 2 bronchial brushings) using the weighted trimmed mean of M-values method from calcNormFactors^29^ in the edgeR^30^ R package to calculate library size factors (**Figure S2**).

### Identification of gene features associated with bronchial dysplasia

For each gene expression dataset (log cpm values for RNA sequencing data or log RMA values for microarray data) used in the training or external testing, the dataset was batch corrected using reference-batch ComBat^31^ to the GSE109743 discovery set using the intersecting set of genes between the two datasets (**Figure S2**). Within each fold of the 5-fold cross validation, prior to gene selection, we retained only genes that were measured across all training samples. For the gene selection, we identified genes associated with dysplasia status (dichotomized as dysplasia or high-grade lesions versus metaplasia, hyperplasia, or normal) using a linear mixed effects model using lmFit^32^ from the limma^33^ R package with smoking status (current or former) and a dataset indicator as covariates and patient as a random effect (included using the limma duplicateCorrelation^34^). Z-score normalized batch corrected gene expression values were used in heatmap visualizations (**Figure 4A**). Within each fold, genes were sorted based on the t-statistic for the dysplasia status, and the top 100 most up- and down-regulated genes were selected. Rows and columns were clustered by the Ward method^35^. We identified significantly enriched pathways among the selected genes using the enrichR^36^ R package, benchmarking against gene sets from Hallmark 2020 (MSigDB^37,38^), KEGG 2021^39,40^, and BioCarta 2016 (accessed through enrichR). For visualization, we focused on the Hallmark 2020 pathways (**Figure 4B**). We also identified significant overlaps between our up- and down-regulated genes and previously defined gene groups from PCA Dataset 1^6,10^,the Colorado SPORE^2^ cohort, and the J.B. Institute cohort using the GeneOverlap^41^ R package (**Figure 4C**).

**Figure 4.**
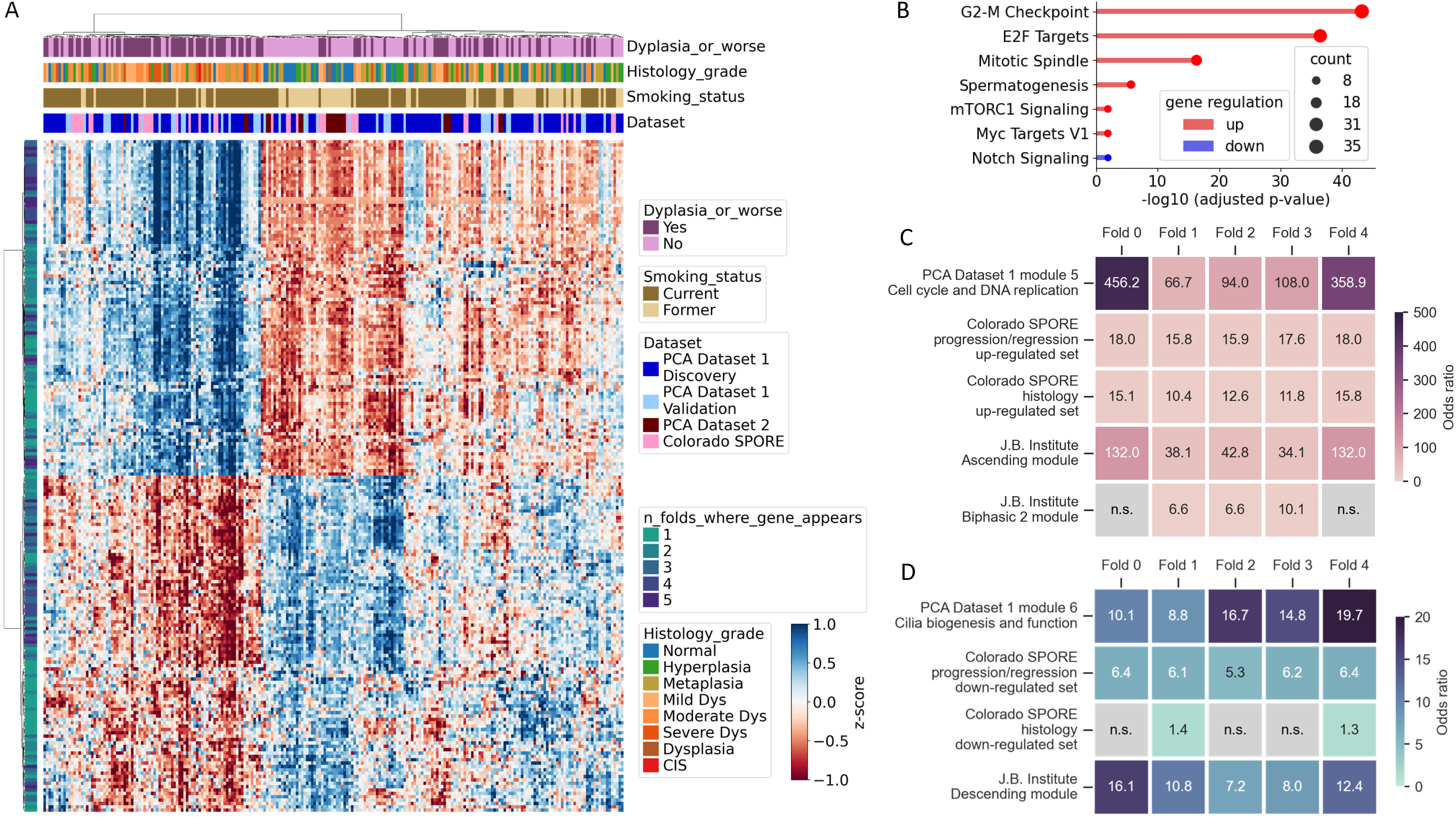
Gene heatmap, pathway analysis, and overlap analysis. **A.** Gene expression (log cpm for RNA-seq and log RMA values for microarray) for the 200 genes from the best-performing fold of the flexible fusion mode is plotted in the heatmap. The heatmap column annotations indicate sample labels (dysplasia or not), histology grade (normal, hyperplasia, metaplasia, mild dysplasia, moderate dysplasia, severe dysplasia, ungraded dysplasia, CIS), smoking status (current or former), and cohort information. The heatmap row annotation shows the number of folds where the gene appeared. Rows and columns were clustered using the Ward method, a hierarchical clustering approach that minimizes the total within- cluster variance. **B.** Gene pathway analysis was implemented on the same set of genes shown in panel **A** against the Hallmark 2020 benchmark from Human Molecular Signatures Database (MSigDB). Enrichment scores (-log 10 (adjusted p-value)) and gene counts were shown for all statistically significant pathways. **C, D.** Gene overlap analysis was conducted for up- (**C**) and down- (**D**) regulated genes, respectively, with pre-defined gene groups from PCA Dataset 1, Colorado SPORE, and J.B. Institute. For each fold, the odds ratios of the fold-specific gene set and pre-defined gene groups were reported for up- or down- regulated genes, respectively.

### Deep learning framework

Our modeling framework consists of three parts (**Figure 1B**): 1) a feature projection module that projects image and gene inputs to feature embeddings, 2) a transformer^42^ encoder that learns the contribution of individual tokens through the self-attention mechanism, and 3) a multilayer perceptron (MLP) head as a classifier. The feature projection module processes input features to generate feature embeddings. It consists of two channels, each with a mask to handle the presence or absence of data during training or inference. The gene channel projects the input gene expression matrix, using selected gene features, into an embedding space through a fully connected network. The image channel, comprising a graph convolutional block, a pooling block with minCUT pooling^43^, and a dense block with layer-wise normalization,^44^ projects input images into the same embedding space. Batch normalization was avoided to ensure compatibility with singleton batches. The projected gene and image features are then passed through a transformer encoder and an MLP head to predict the probability of bronchial dysplasia or worse histology on a per-sample basis. As such, our model builds upon the Vision Transformer (ViT)^45^ architecture by incorporating parallel channels to project inputs from different feature spaces into a unified embedding space, enabling effective multimodal data integration. It is designed to handle data variability, offering flexibility to accommodate incomplete features within a dataset and differences in data modalities across samples. Additionally, by leveraging a graph-based architecture, our model achieves greater computational efficiency, as the adjacency matrix in the graph convolutional layers implicitly encodes spatial relationships, serving a role analogous to positional encoding in vision transformers while preserving both local and global context among image tokens.

### Experimental design and performance metrics

The multimodal transformer was trained to perform binary classification (normal/hyperplasia/metaplasia versus dysplasia/CIS/SCC) (**Figure 1C**). Training and internal testing with 5-fold cross validation were performed using five data cohorts with patient-level stratified sampling within each cohort containing WSIs or gene expression (GE) data or both: PCA Dataset 1 Discovery set (WSI and GE), PCA Dataset 1 Validation set (WSI and GE), PCA Dataset 2 (WSI and GE), Pennycuick (WSI), and Colorado SPORE (GE). Five models were trained and tested (**Figure S3**): 1) single-modality models (**Figure 2A**): Model *a* (trained on WSI, tested on WSI) and Model *c* (trained on GE, tested on GE); 2) fusion models (**Figure 2A**): Model *b* (trained on WSI and GE, tested on WSI) and Model *d* (trained on WSI and GE, tested on GE); 3) the flexible fusion model (trained on samples regardless of the number of data modalities) (**Figure 3A**). Model *b* and Model *d* were trained on the same samples, but different model notation (*b*, *d*) was used to highlight differences in the model testing. Average performance metrics, including AUROC and AUPR (area under receiver operating characteristic and precision-recall curves), accuracy, precision, sensitivity/recall, and specificity, of all models were reported along with standard deviation across folds. Probability thresholds for obtaining accuracies were optimized on internal testing samples with the highest internal accuracy and were applied to external samples for the external accuracy as well. Precision is a fraction of true positives over the total number of positive predictions, sensitivity/recall measures the proportion of true positives that are correctly identified, and specificity measures the proportion of true negatives that are correctly identified. The statistical significance of AUROC differences was assessed using DeLong’s test^46^. The Mann-Whitney U test^47^ was conducted on top two principal components of z-score normalized feature embeddings from the model’s penultimate layer, with p-values indicating the statistical significance of PC value distributions between samples with histological grades of dysplasia or worse versus normal, hyperplasia, and metaplasia (non-dysplasia).

To ensure consistent model performance, external testing was conducted on DECAMP-1 and J.B. Institute cohorts. Results were reported separately by the testing sample type: 1) WSI samples from DECAMP-1, 2) GE samples from DECAMP-1 and J.B. Institute, 3) WSI + GE samples from DECAMP- 1 (**Figure 3A**). We also tested the flexible fusion model’s predictive performance on PCA bronchial brushing GE samples. Average performance metrics were reported along with standard deviation across folds. Model weights of the fold with the highest AUROC score on the external cohorts were used for post hoc analysis. The 128-dimensional features from the penultimate layer of the model were extracted for principal component analysis^48,49^ to visualize the relationship between histologic grades and WSI or GE features.

Optimal hyperparameters for the transformer configuration included a hidden dimension of 128, a graph block with three graph convolution layers, a minCUT pooling^50^ configuration of 50 clusters, and one encoder block in the transformer encoder. Four heads were used for multi-head attention training. To avoid overfitting, dropouts with a rate of 0.2 were used in feed-forward blocks, and binary cross-entropy loss to enhance robustness. For Models *a, b*, *d*, training was completed over 30 epochs, a 0.00005 initial learning rate, and a step scheduler with 0.5 decay at epoch 15. For Model *c* and the flexible fusion model, training was completed over 50 epochs, a 0.0001 initial learning rate, and a step scheduler with 0.5 decay at epoch 15 and 30. All models were trained on 8-sample minibatches and used the Adam optimizer^51^ for expedited convergence. All the experiments were performed on a single GeForce RTX 3090 24 Gb workstation.

### Data and code availability

Data will be available in the public domain upon publication of the manuscript. Computer scripts and manuals will be made available on GitHub upon the publication of the manuscript.

## Results

In this study, we used whole slides images (WSIs) of H&E stained sections and/or bulk gene expression (GE) data from endobronchial biopsies obtained from high-risk patients for developing lung cancer from multiple cohorts (**Figure 1A**). Summary statistics and clinical features including gender, age, race, and smoking status are reported in **Table 1**. For all cohorts, over half of the subjects are current versus former or never smokers. The percentage of subjects with a histological grade of dysplasia or higher varied across the cohorts, ranging from 13.9% (DECAMP-1) to 100% (Jules Bordet Institute). The gender ratio (male over female) also differed greatly across the cohorts, ranging from a balanced rate of 53.5% (PCA Dataset 1) to a highly imbalanced rate of 15.3% (DECAMP-1). Leveraging samples from diverse populations, we trained (**Figure 1B**) and evaluated five models, defined by the data used for training and testing: two single modality models (Model *a* and Model *c*) trained and tested on samples with only WSI or only GE, two fusion models (Model *b* and Model *d*) trained on samples with both WSI and GE and tested on samples with only WSI or only GE, and a flexible fusion model trained on samples with either WSI or GE or both and tested on samples with WSI only, GE only, or both (**Figure 1C**).

### Fusion model has higher performance than single modality models in predicting bronchial dysplasia

Our transformer framework demonstrated improved performance utilizing two data modalities (GE and WSI) over a single modality (GE or WSI) and robustness across the five-fold cross-validation indicated by concordance in the true and predicted classes across different folds (**Figure 2A**). As indicated by the receiver operating characteristic (ROC) curves on internal testing samples (**Figure 2B**), combining GE and WSI during training resulted in area under ROC curve (AUROC) of 0.766±0.022 on WSI samples (Model *b*), with an increase of 0.064 compared to the AUROC of 0.702±0.027 when using WSI only (Model *a*) (p<0.0001). Similarly, using both GE and WSI for training achieved the AUROC of 0.876±0.021 on GE samples (Model *d*), improving the AUROC of 0.823±0.028 when using GE only (Model *c*) by 0.053 (p<0.0001). The area under the PR curve (AUPR) scores from the precision-recall (PR) curves showed the same improvements as AUROCs (**Figure 2B**). The external testing performance (**Figure 2C**) was consistent with above findings from internal testing, with the AUROC improved from 0.690±0.027 (Model *a*) to 0.761±0.015 (Model *b*) (p<0.0001), and from 0.816±0.032 (Model *c*) to 0.890±0.023 (Model *d*) (p<0.0001). In concordance with these results, samples with a histologic grade of dysplasia or worse had higher predicted probabilities compared to non-dysplasia samples in Model *b* vs. Model *a* (**Figure S5A**) and in Model *d* vs. Model *c* (**Figures S5B & S5C**) across external testing samples.

While the transformer framework was trained to distinguish samples with histological grades of dysplasia or worse versus normal, hyperplasia, and metaplasia, we observed improved stratification between histological grades within and between the two prediction classes using dual over single data modalities. The sample prediction probabilities of Model *a*, tested on DECAMP-1 WSIs, were different between dysplasia versus normal, hyperplasia, and metaplasia (all comparisons p<0.0001, **Figure S4A**), while Model *b* probabilities were also different between normal and metaplasia (p<0.05, **Figure S4A**). Feature embeddings from the penultimate layer of each model were also visualized, where z-score normalized high-dimensional GE or WSI features were reduced to the top two principal components (PCs) that associate with the most important patterns the model utilized for classification. The separation between dysplasia versus non-dysplasia among DECAMP-1 WSI samples is more pronounced in the PC plot of Model *b* (PC1: p<0.01, PC2: p<0.05) versus Model *a* (PC1: p<0.001, PC2: n.s.) (**Figure 2H**), evaluated by the Mann-Whitney U test on the distribution of PC values between samples with histological grades of dysplasia or worse versus normal, hyperplasia, and metaplasia. For GE samples, both Model *c* and Model *d* performed well on the DECAMP-1 cohort with significant probability differences between all histology grades (all comparisons p<0.01, **Figure S4A**). On the J.B. Institute cohort, Model *c* probabilities were not different between dysplasia and CIS or CIS and SCC (**Figure S4A**), whereas Model *d* (**Figure S4A**) had probabilities that were different between CIS against SCC (p<0.01), but not between CIS and dysplasia.

The PC plot of Model *d* (PC1: p<0.0001, PC2: p<0.05) compared to Model *c* (PC1: p<0.0001, PC2: n.s.) (**Figure 2H**) indicated better separation between dysplasia versus non-dysplasia among DECAMP-1 and J.B. Institute GE samples.

### Flexible fusion model has the highest performance predicting bronchial dysplasia

The flexible fusion model extended the fusion model (Models *b* and *d*, **Figure 2**) by relaxing the requirement of paired WSI and GE data and allowing the training data to include either GE or WSI or both modalities (**Figure 3**). The internal testing AUROC on WSIs was 0.829±0.029, an improvement of 0.063 compared to Model *b* (p<0.001). Similarly, the internal testing AUROC on GE was 0.915±0.027, an improvement of 0.048 compared to Model *d* (p<0.01) (**Figure 3B**). In the external testing data, the AUROC on WSIs was 0.809±0.036, an improvement of 0.048 compared to Model *b* (p<0.05) and the AUROC on GE was 0.903±0.022, an improvement of 0.013 compared to Model *d* (p<0.01) (**Figure 3C**). In addition, the flexible fusion model was tested on samples with both WSI and GE and resulted in AUROC of 0.928±0.034 and 0.908±0.030 on internal (**Figure 3B**) and external (**Figure 3C**) samples, respectively, with improvement from using GE only by 0.013 (internal, p<0.05) and 0.005 (external, p<0.05), respectively. The precision-recall (PR) curves showed a consistent pattern with ROC curves, where the flexible fusion model had higher PR values tested on WSIs or GE compared to Model *b* and Model *d*, respectively. The highest PR and ROC curves were achieved when the test data contained both WSI and GE (**Figures 3B & 3C**). The comparison of models indicated improved performance using a larger training dataset even if each sample did not contain all the data modalities, and as expected, inclusion of all data modalities during inference improved performance (**Figure S3**).

The flexible fusion model probabilities were different between histological grades within and between the two prediction classes. On the DECAMP-1 cohort, utilizing both WSI and GE for inference resulted in different sample prediction probabilities between all histology grades (all comparisons p<0.01, **Figure S4B**). On the GE samples of both the DECAMP-1 and the J.B. Institute cohort, the prediction probabilities were also different between all histological grades (all comparisons p<0.01, **Figure S4B**). The PC plot on WSI samples in the flexible fusion model (PC1: p<0.001, PC2: p<0.05) improved separation between dysplasia and non-dysplasia samples compared to the fusion model (Model *b*). This improvement was also seen on GE samples in the flexible fusion model (PC1: p<0.0001, PC2: p<0.01) (**Figure 3G**) compared to the fusion model (Model *d*).

### Gene features used to predict dysplasia status are associated with proliferation pathways

Gene expression alterations associated with dysplasia status were chosen using four datasets in the flexible fusion model. A meta-analysis across these four datasets hasn’t been conducted, so we wanted to examine the biology of the gene features as well as their relationships to previously identified gene signatures associated with histologic severity. A heatmap of the genes in the best performing fold (**Figure 4A**) shows z-score gene expression values where 4.36% of genes were chosen across all folds and 20.87% of genes were chosen by at least 3 folds. Since the genes were chosen using a linear regression model, larger versus smaller datasets may have a greater influence despite including dataset as a covariate, however, there was no significant difference found in model performance among training cohorts (p>0.05 from the Kruskal-Wallis test^49^ on bootstrapped AUROCs). The up-regulated genes in samples with dysplasia or worse histology were enriched in six pathways (from Hallmark 2020^37,38^) associated with cell cycle, E2F targets and mTORC1 signaling and down-regulated genes were enriched in Notch signaling (**Figure 4B**). Sixty-five percent of genes responsible for the significant enrichment of the up-regulated pathways were associated with more than one pathway, including AURKA (5 out of the 6 pathways) and BUB1, PLK1, KIF2C, CCNB2 (4 out of the 6 pathways). PLK1 (polo-like kinase) was previously associated with persistent bronchial dysplasia and inhibition of PLK1 induced apoptosis and decreased proliferation in cultured cells derived from these lesions^2^. The genes also significantly overlapped with genes previously derived from PCA Dataset 1^6,10^, the Colorado SPORE^2^ cohort, and the J.B. Institute^7^ cohort. Up-regulated genes significantly overlapped with the gene co-expression module (module 5) associated with cell cycle and DNA replication from PCA Dataset 1, the up-regulated genes in progressive versus regressive samples from the Colorado SPORE cohort, the up-regulated genes with increasing histologic severity from the Colorado SPORE cohort, and the ascending module associated the proliferation from the J.B. Institute cohort across all folds (**Figure 4C**). Besides, three out of five folds significantly overlapped with the biphasic 2 module from the J.B. Institute cohort. Down-regulated genes significantly overlapped with the gene co-expression module (module 6) associated with cilia biogenesis and function from PCA Dataset 1, the down-regulated genes in progressive versus regressive samples from the Colorado SPORE cohort, and the descending module linked to DNA damage response downregulation from the J.B. Institute cohort across all folds (**Figure 4D**). There was also significant overlap in two out of five folds with the down-regulated genes associated with increasing histologic severity from the Colorado SPORE cohort.

### Flexible fusion model predicts presence of bronchial dysplasia using normal appearing bronchial brushings

We have previously shown that gene expression derived from cells collected from a normal appearing area of right or left mainstem bronchus via brushing can be leveraged to predict the presence of lung squamous premalignant lesions^6,52^, lung cancer^53–56^, and chronic obstructive pulmonary disease^57^. Based on these studies, we sought to determine if the flexible fusion model, trained on endobronchial biopsies, would be able to discriminate between airways where a bronchial dysplasia or worse lesion was sampled versus airways containing no evidence of dysplasia. The flexible fusion model tested on 174 bronchial brushing samples (137 from PCA Dataset 1, 37 from PCA Dataset 2, **Figure 5B**) achieved AUROC of 0.721±0.015 (**Figure 5E**) and AUPR of 0.741±0.014. The flexible fusion model showed marginal improvement in AUROC from model *c* by 0.018 (p>0.05) and model *d* by 0.008 (p>0.05) (**Figures 5C & 5D**), and therefore, it is not clear whether the inclusion of the WSIs enhances model performance in other sample types with only GE (e.g. bronchial brushings versus endobronchial biopsies). A Mann-Whitney U test showed significant predicted probability differences between samples with histological grades of dysplasia or worse versus normal, hyperplasia, and metaplasia (p<0.0001).

**Figure 5.**
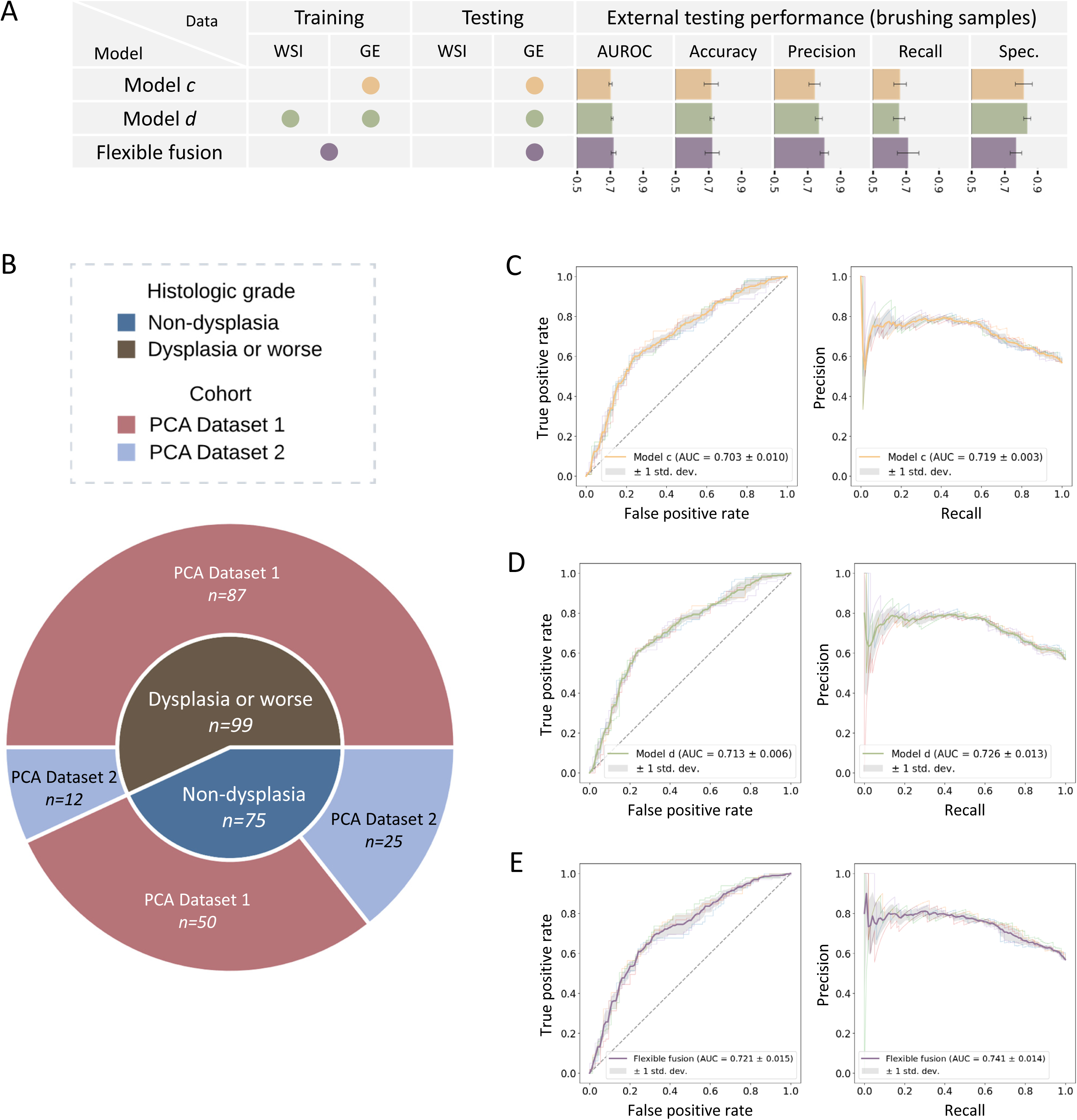
Classification performance on brushing samples. **A.** Brushing samples were tested with Model *c*, Model *d*, and the flexible fusion model. The same performance metrics as Figure 2 were visually reported in the bar charts. **B.** Out of the 174 bronchial brushing samples, 137 are from PCA Dataset 1, and 37 are from PCA Dataset 2, with 99 being dysplasia or worse and 75 being non-dysplasia (normal, hyperplasia, or metaplasia). **C, D, E.** Receiver operating characteristic (ROC) and precision- recall (PR) curves showcase the binary classification performance of Model *c* (**C**), Model *d* (**D**), and the flexible fusion model (**E**). The area under the curve (AUC) score was reported with standard deviation across five folds.

## Discussion

In this work, we developed a multi-modal transformer framework designed to distinguish between endobronchial biopsies with bronchial dysplasia or worse histology versus biopsies with normal, hyperplasia, or metaplasia histology by integrating H&E-stained WSIs and bulk GE data. Despite knowing that histopathologic and transcriptomic alterations are both associated with severity and progression of PMLs^6,13,52^, gene expression data is often not available from biopsies outside of research studies. Also, among research studies, the study of PMLs across different tissue types by initiatives like the National Cancer Institute-funded Human Tumor Atlas Network (HTAN) has profiled lesions using different methodologies. We sought to develop a generalizable framework to use samples with one or more data modalities during training or inference. Inspired by the recent work on pan-cancer survival analysis using WSIs and molecular (gene expression, copy number alterations, somatic mutations) features^58^ and non-small cell lung cancer survival analysis using WSIs and bulk gene expression data^59^, our multi-modal transformer aligned image-based morphologic and gene expression features to learn a shared complementary representation of premalignant biology. The framework enhanced single data modality prediction through training on both data modalities and it may enhance our ability to assess histologic severity and longitudinally monitor changes in serial samples collected over time.

Our study utilized several published (PCA Dataset 1^6,10,13^, J.B. Institute^7^, Colorado SPORE^2^, Pennycuick^8^) and unpublished (PCA Dataset 2, DECAMP-1) cohorts with data containing H&E WSI and/or bulk GE of endobronchial biopsies to train and test models. We compared model performance between models trained on WSI or GE to a model trained on paired WSI and GE data from the same samples and showed that the model trained on both modalities outperformed the single modality models in both the internal and external testing sets. The external testing WSIs from DECAMP-1 indicated significantly higher performance (AUROC) in Model *b* (WSI+GE) compared to Model *a* (WSI). Similarly, the external testing GE from DECAMP-1 and J.B. Institute indicated significantly higher performance (AUROC) in Model *d* (WSI+GE) versus Model *c* (GE). Our results suggest that leveraging data from multiple modalities during training can enhance prediction performance when only a single data modality is available. This observation indicates that the model learns a shared and complementary representation of lung squamous premalignant biology by leveraging cell morphology and spatial organization from H&Es and gene expression measurements from RNA-seq or microarrays. Given the higher performance of the model trained on both data modalities, we built a flexible fusion model that allowed the inclusion of non-paired data (WSI or GE only) during training. The flexible fusion model showed significant improvement over the fusion model trained on paired WSI and GE data during external single modality testing on WSIs and GE. The flexible fusion model tested on both data modalities had the highest performance, but the performance was most improved when testing using WSIs only, potentially implying that the model could be enhanced with additional WSI training data.

Our multi-modal transformer was initially trained on binary labels to distinguish PMLs that are likely dysplasia or a worse histology from those that are lower histologic grade. However, post hoc analysis examining the predicted probabilities revealed that the models were able to stratify histology grades within the binary categories. These results suggest that the features learned during training captured more nuanced information associated with histologic severity. The flexible fusion model showed significant differences between the prediction probabilities of all histologic grades including normal, hyperplasia, metaplasia, mild dysplasia, moderate dysplasia, severe dysplasia, CIS, and SCC. The framework provided additional insights into more detailed histologic grades without requiring extensive retraining on finely graded labels, suggesting its potential to be repurposed or refined for more granular classification tasks. Additionally, the framework may have utility in providing a continuous metric for longitudinal monitoring of lesions, and future work could assess how changes in prediction probabilities overtime are related to lesion progression to SCC. The gene features selected across multiple cross-validation folds and datasets may indicate reproducible gene expression changes associated with histologic severity that could be potential interception targets. For example, Merrick *et al*.^2^ showed decreased proliferation and increased apoptosis treating cells from lesions with persistent bronchial dysplasia with an inhibitor of PLK1, one of the genes selected by our model. In future work, it will be interesting to assess the efficacy of other inhibitors to genes like AURKA and BUB1 that were identified by our model.

Prior work has shown that a subset of gene expression changes associated with higher-grade PMLs can be reflected in normal appearing airway epithelium^6,52^, so we wanted to test the flexible fusion model’s ability to predict the presence of bronchial dysplasia in the lung using GE from bronchial brushings collected from normal appearing areas of the mainstem bronchus. The flexible fusion model had an AUROC of 0.721±0.015 which was lower than model *c*, *d*, or the flexible fusion model tested on GE from endobronchial biopsies; however, the prediction probabilities were significantly different between brushes where the worst histology sampled (via biopsy during the procedure) was dysplasia versus normal, hyperplasia, or metaplasia. While this model may have utility in predicting other sample types from the airway field, the performance of the flexible fusion or fusion model was similar to the GE only model indicating that training on both WSIs and GE may not provide a performance benefit in other sample types (brushings) collected from lung sites far from the site of the PML.

Our models were trained using pathologist-defined histologic grades, but no centralized review was conducted for the H&E slides. This is a limitation given the well-documented intra- and inter-observer variability in grading of PMLs, which are not routinely evaluated in clinical practice. Also, the pathologists that assessed the flexible fusion model training versus external testing samples were different, so we are unable to assess in external testing data the impact of pathologist bias. Despite this limitation, a key advantage of our approach is that it can enable consistent assessment of PMLs across institutions requiring centralized review, a process that is time-consuming and resource intensive. Analysis of multi-omic data from lung PMLs^2,5,7,8,13^ indicates that there are molecular changes in the immune microenvironment and within the epithelium that contribute to lesion severity and risk of progression that are not easily identifiable in H&E slides. Future work may benefit from training using a label that is created based on a consensus of pathologic and molecular information to achieve better dichotomization. Additionally, the WSI data used in this study was obtained from forceps endobronchial biopsies, that are small and often contain tissue fragments, potentially limiting the model’s ability to learn global and contextual features or regional information. As a result, future extensions could involve training the model using lung resection samples where lung PMLs are present within the tumor margins and integrating additional data modalities such as single cell sequencing and spatial transcriptomics as it becomes available. These comprehensive approaches could provide deeper molecular insights into dysplastic patterns that are more aggressive and prone to progression invasive carcinoma. Additionally, while we used CTransPath^16^ to extract image-based feature vectors, future work should explore leveraging alternative vision foundation models, including those pre-trained on diverse pathology datasets. Deep learning models such as GenePT^60^, which integrates genomic and transcriptomic context from histopathologic data, represent a promising direction for replacing the modeling approach currently used in our pipeline.

In summary, our multi-modal transformer can efficiently process two data modalities, WSIs and bulk GE data, obtained from endobronchial biopsies and predict bronchial dysplasia or worse histology. The model outperformed models trained on a single data modality and enabled the inclusion of samples with one or both modalities during training and/or testing. As more methods are used to measure the biology of PMLs, this approach increases the flexibility, scalability, and real-world applicability of disease severity assessment that may better risk stratify PMLs or longitudinally monitor PMLs even when only routine histology data is accessible.

## Disclaimer

The content is solely the responsibility of the authors and does not necessarily represent the official views of the National Institutes of Health.

## Data Availability

All data produced in the present study are available upon reasonable request to the authors.

## Acknowledgement

We would like to thank the Lung PCA Consortium and the DECAMP Consortium for their invaluable role in consenting study participants and collecting, sharing, and profiling biospecimens utilized in this manuscript. We want to specifically acknowledge the contributions of the Lung Cancer Data and Biospecimen Research Teams at Roswell Park (Emily Siedlecki, Kelsey Simon, Britt Holdaway-Kenney, Debra Bradley, and Lindsey Steinwandel), University College London (Kate Gowers, Kate Otter, and Lukas Kalinke), and University of Colorado (Dan Merrick, Hannah Schumann, Adrie Bokhoven, and Carsten Goerg) to the Lung PCA.

---

**Figure S1.**
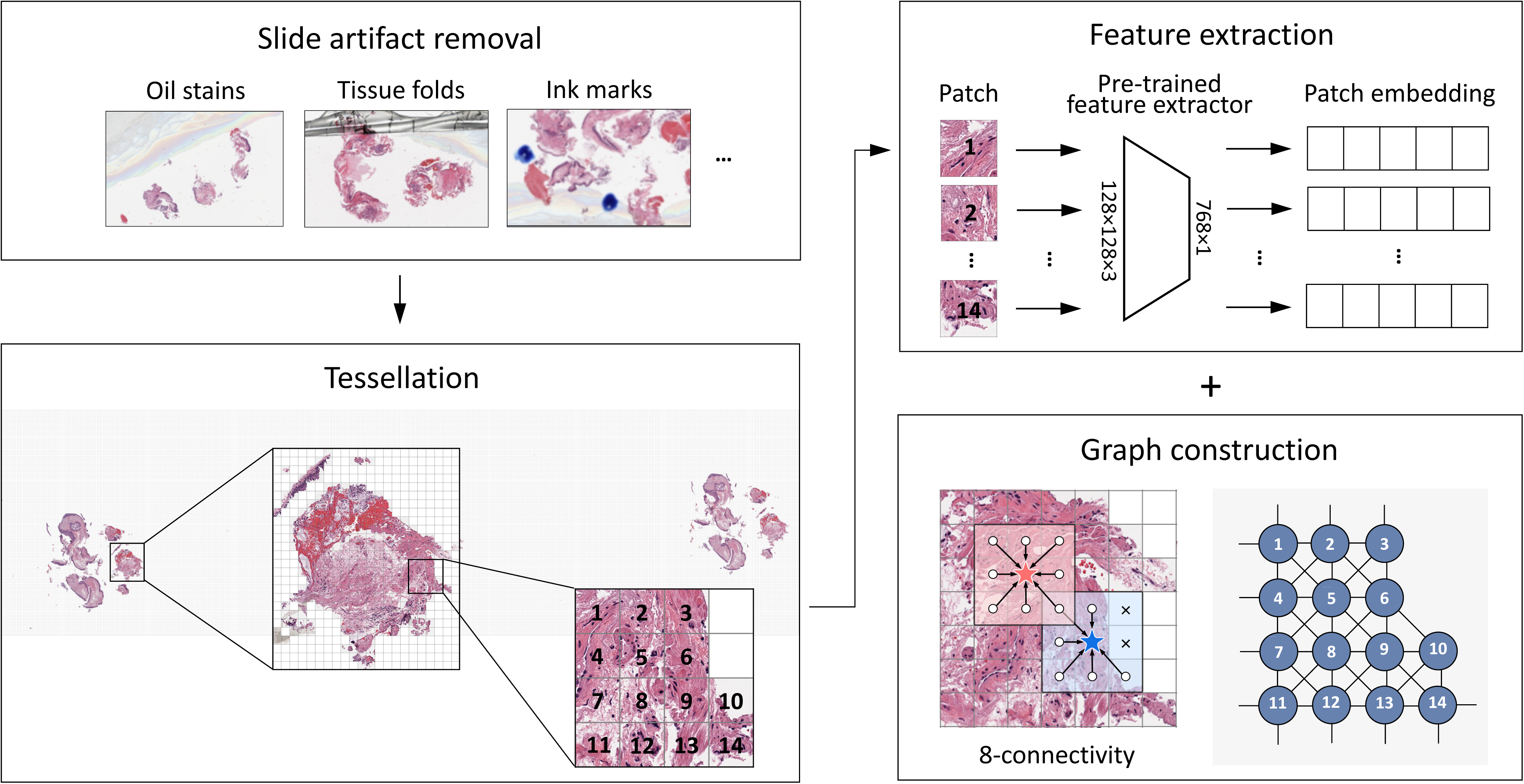
Whole slide images pre-processing. WSIs were processed by a pipeline that separated tissues from backgrounds and artifacts, meanwhile generating squared image patches and constructing them into an undirected graph. Patch embeddings were computed using cTransPath and used as node features in the graph. Edges were built upon 8-connectivity.

**Figure S2.**
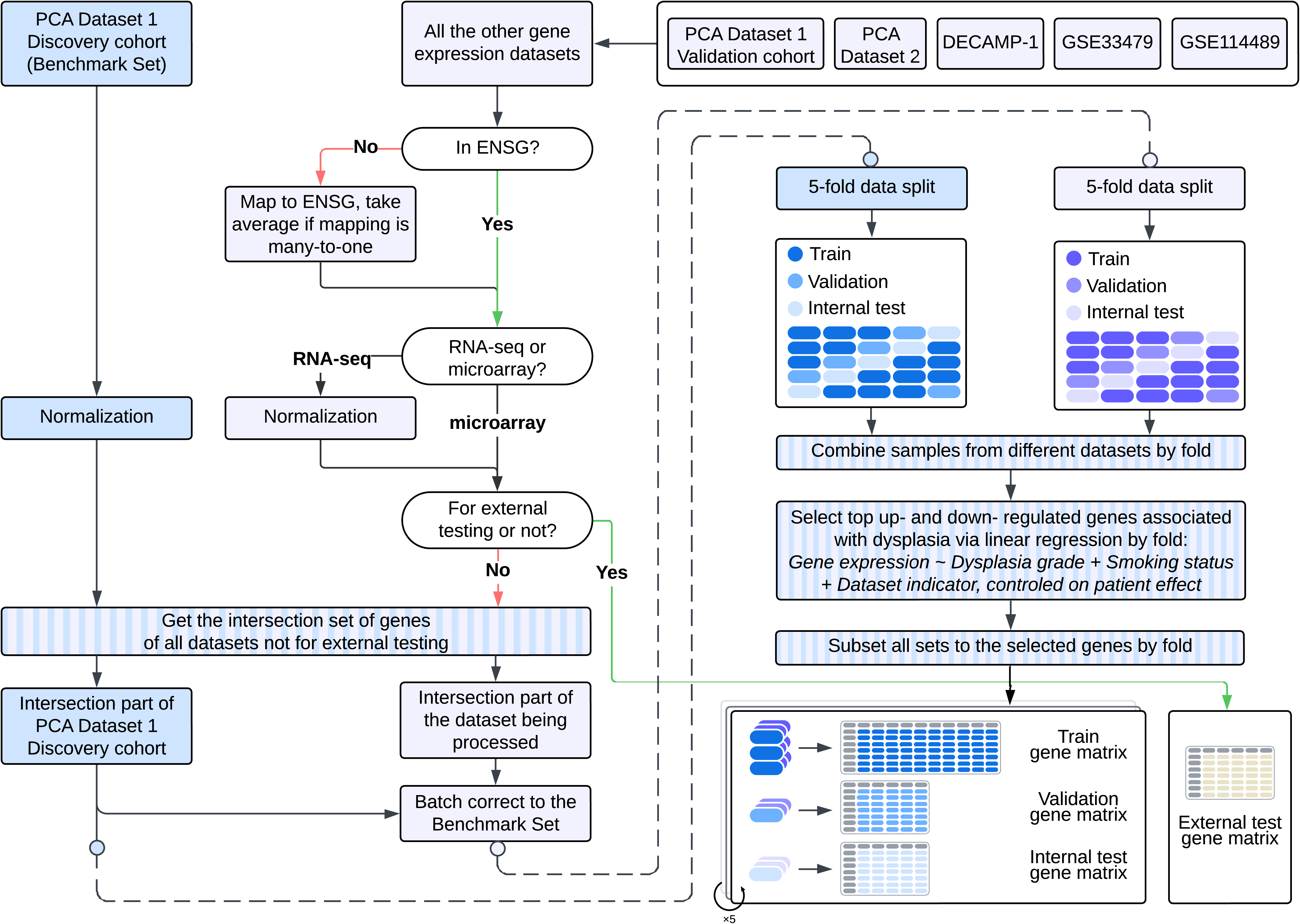
Gene expression data pre-processing. All datasets for training and internal testing were batch corrected to PCA Dataset 1 discovery cohort on the intersection set. Ensembl transcript IDs were mapped to ensemble gene IDs before intersection. Patient-level stratified sampling (**Figure S3**) was conducted within each training dataset. Linear regression was implemented on all training samples per fold to derive the set of genes associated with dysplasia based on ranking of up-regulated and down- regulated genes. Top 100 up- and down- regulated genes were selected per fold. Datasets for external testing were also batch corrected to PCA Dataset 1 discovery cohort.

**Figure S3.**
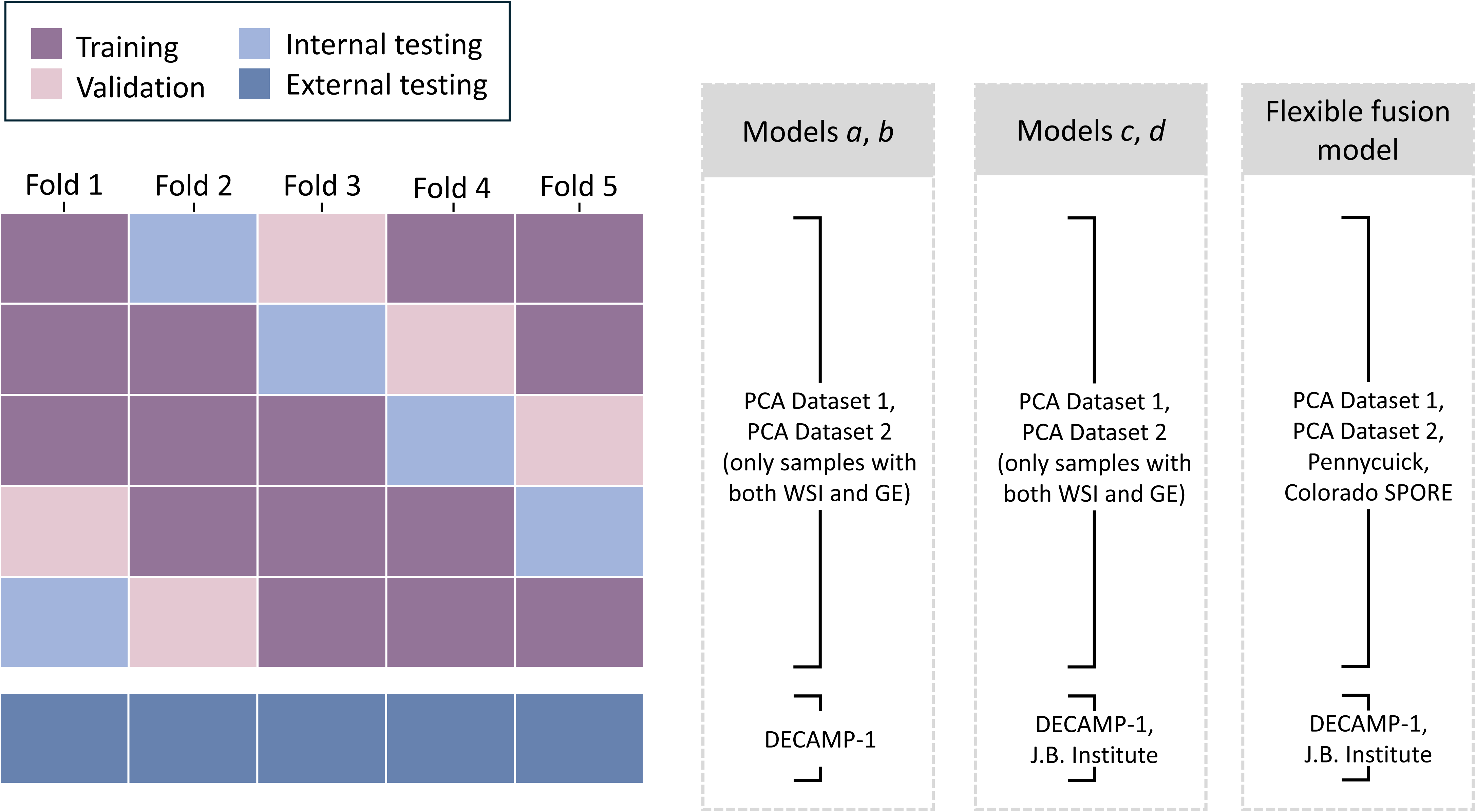
Data split and cross-validation. Models were trained and internally tested on PCA Dataset 1 & 2, Pennycuick, and Colorado SPORE cohorts, using samples with WSI (Model *a*), samples with GE (Model *c*), samples with both WSI and GE (Model *b* and Model *d*), or all samples (flexible fusion model). Qualified samples from the above datasets were randomly divided into five folds through stratified split at the patient level. Models were externally tested on DECAMP-1 and Jules Bordet Institute cohorts.

**Figure S4.**
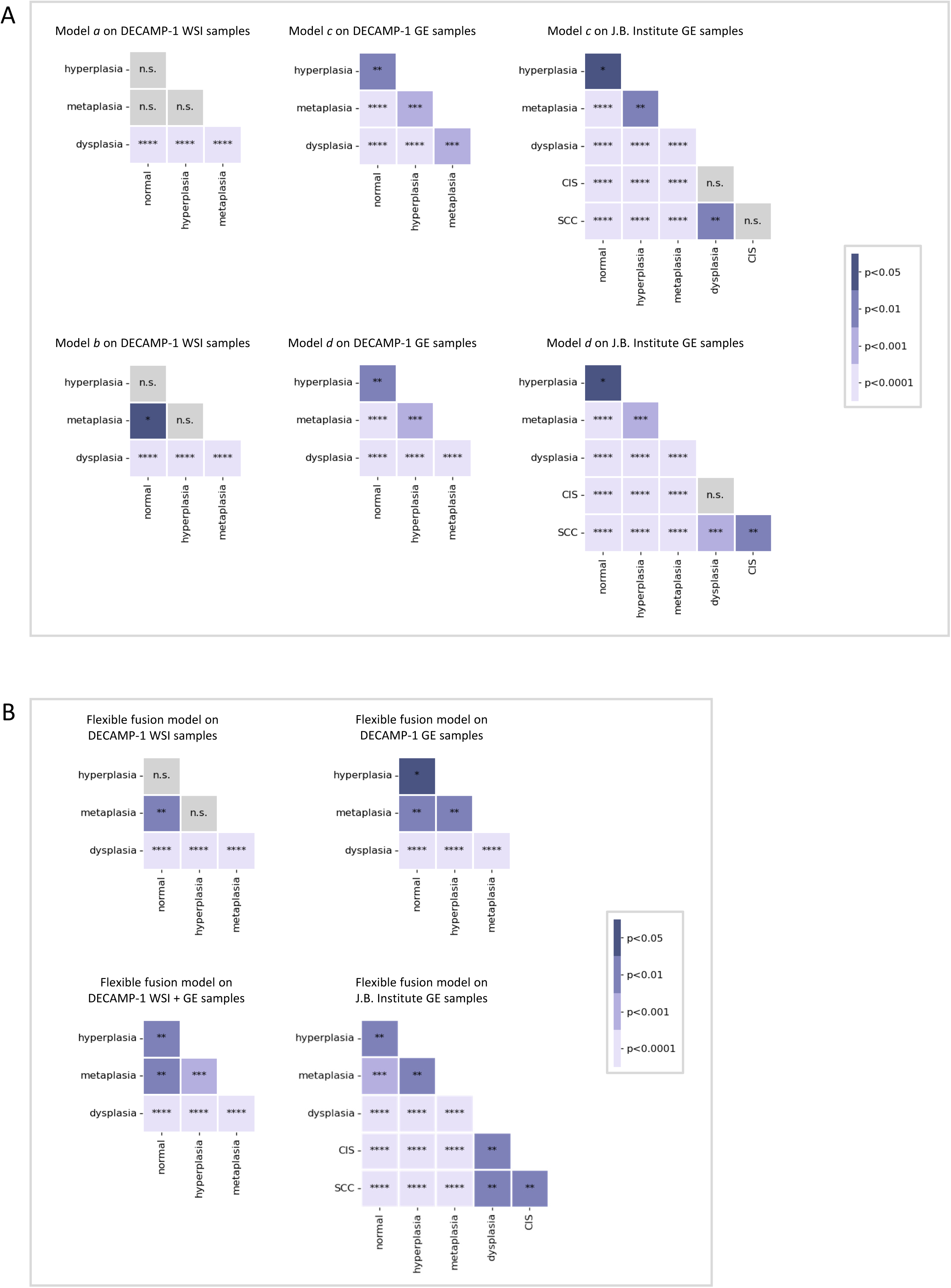
Heatmap of statistical significance of boxplots from Figure 2 and Figure 3. **A.** P-values of all histologic grade pairs in boxplots **Figures 2D-2G**. **B.** P-values of all histologic grade pairs in boxplots **Figures 3D-3F**. Significance levels are denoted as * for p<0.05; ** for p<0.01; *** for p<0.001; and **** for p<0.0001.

**Figure S5.**
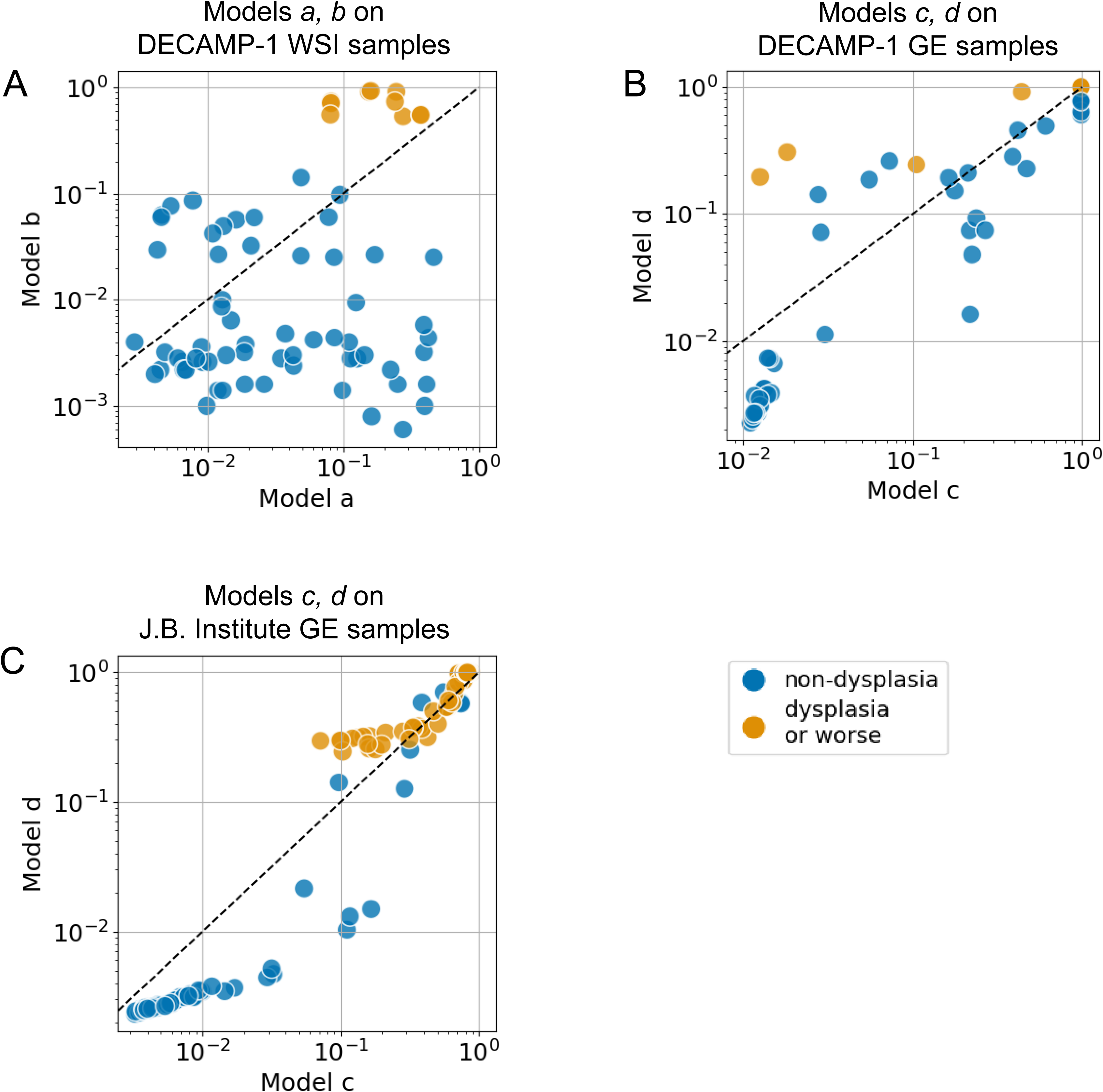
Scatter plot of external testing sample prediction probabilities. Categorized by histologic grades, sample prediction probabilities were plotted for Model *a* against Model *b* on DECAMP-1 WSI samples (**A**), Model *c* against Model *d* on DECAMP-1 GE samples (**B**), and Model *c* against Model *d* on J.B. Institute GE samples (**C**).

**Figure S6.**
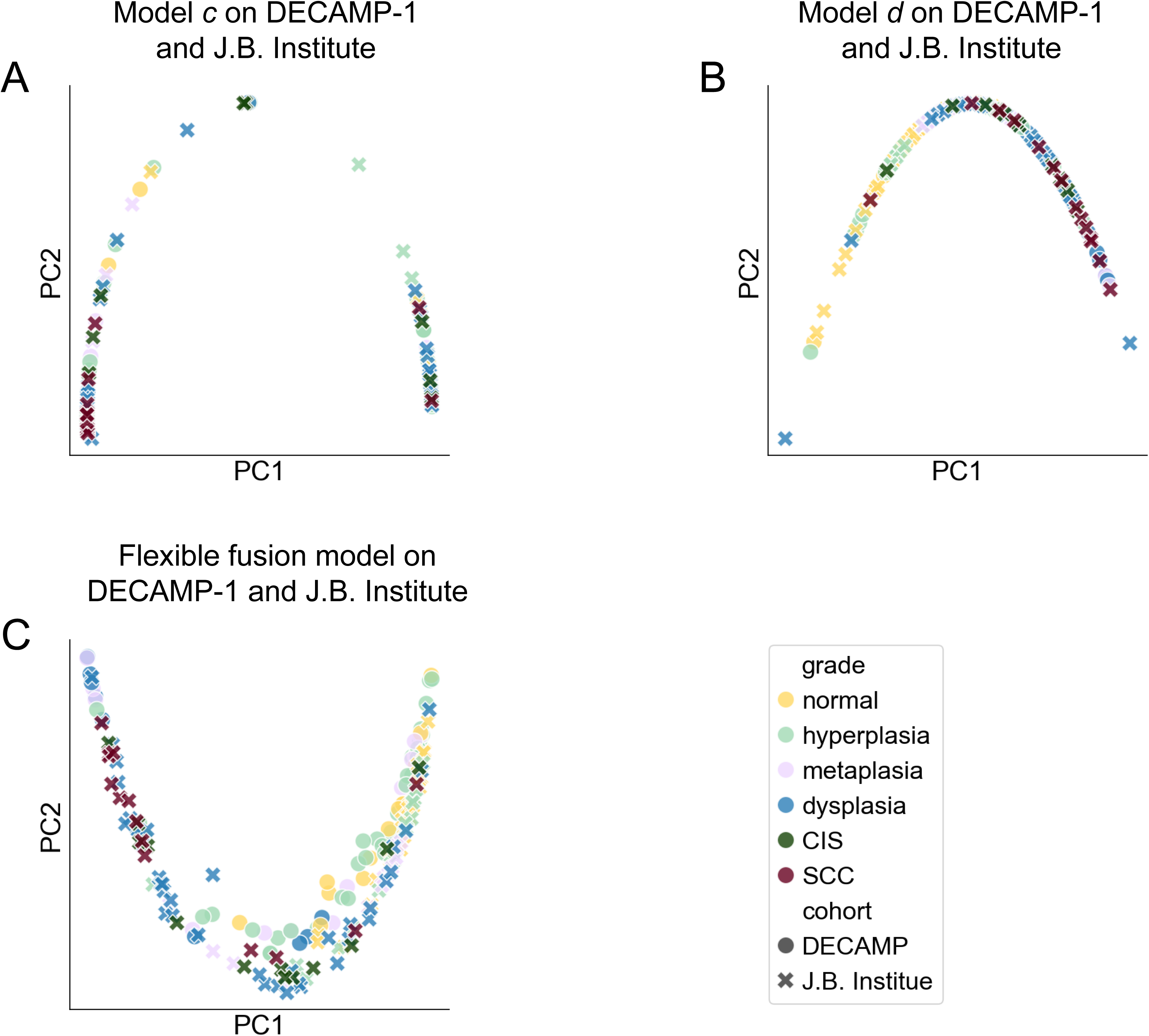
PC plots on external testing samples by cohort (extended from Figure 2H and Figure 3G). **A.** Model *c* on DECAMP-1 and J.B. Institute GE samples. **B**. Model *d* on DECAMP-1 and J.B. Institute GE samples. **C**. The flexible fusion model on DECAMP-1 and J.B. Institute GE samples.

**Figure S7.**
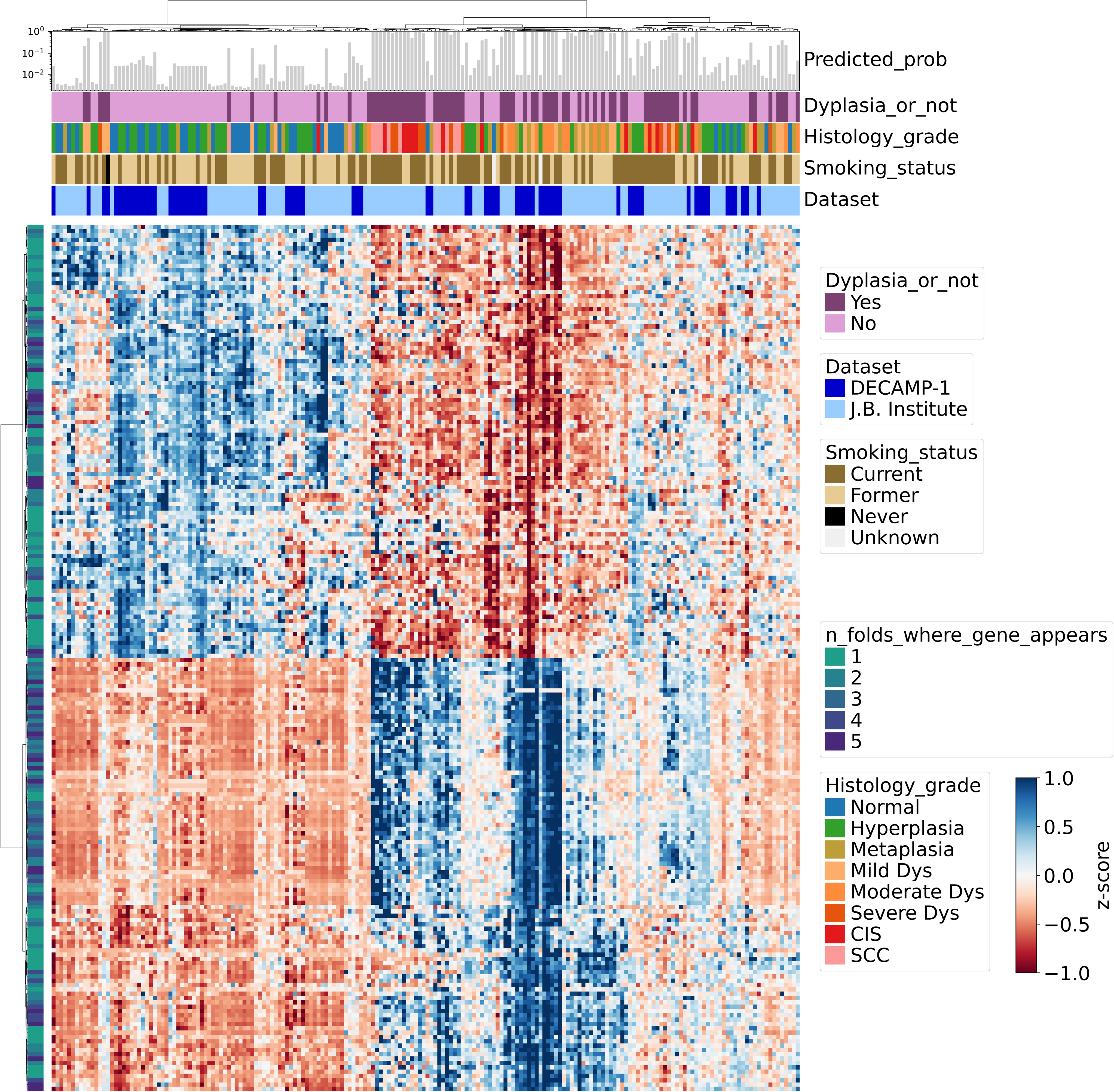
The gene heatmap of external testing samples. Displayed genes are from the intersection of external testing samples and all training samples of the flexible fusion model. Top annotations indicate predicted probabilities, sample labels (dysplasia or not), histology grades (normal, hyperplasia, metaplasia, mild dysplasia, moderate dysplasia, severe dysplasia, ungraded dysplasia, CIS, SCC), smoking status (current, former, never, or unknown), and cohort information (DECAMP-1 or J.B. Institute). The left annotation shows the number of folds where a gene appears. Rows and columns were clustered using the Ward’s method.

